# Early Short Course Corticosteroids in Hospitalized Patients with COVID-19

**DOI:** 10.1101/2020.05.04.20074609

**Authors:** Raef Fadel, Austin R. Morrison, Amit Vahia, Zachary R. Smith, Zohra Chaudhry, Pallavi Bhargava, Joseph Miller, Rachel M. Kenney, George Alangaden, Mayur S. Ramesh, Henry Ford COVID-19 Management Task Force

## Abstract

**Background:** There is no proven antiviral or immunomodulatory therapy for COVID-19. The disease progression associated with the pro-inflammatory host response prompted us to examine the role of early corticosteroid therapy in patients with moderate to severe COVID-19.

**Methods:** We conducted a single pre-test, single post-test quasi-experiment in a multi-center health system in Michigan from March 12 to March 27, 2020. Adult patients with confirmed moderate to severe COVID were included. A protocol was implemented on March 20, 2020 using early, short-course, methylprednisolone 0.5 to 1 mg/kg/day divided in 2 intravenous doses for 3 days. Outcomes of pre and post-corticosteroid groups were evaluated. A composite endpoint of escalation of care from ward to ICU, new requirement for mechanical ventilation, and mortality was the primary outcome measure. All patients had at least 14 days of follow-up.

**Results:** We analyzed 213 eligible subjects, 81 (38%) and 132 (62%) in pre-and post-corticosteroid groups, respectively. The composite endpoint occurred at a significantly lower rate in post-corticosteroid group compared to pre-corticosteroid group (34.9% vs. 54.3%, p=0.005). This treatment effect was observed within each individual component of the composite endpoint. Significant reduction in median hospital length of stay was observed in the post-corticosteroid group (8 vs. 5 days, p < 0.001). Multivariate regression analysis demonstrated an independent reduction in the composite endpoint at 14-days controlling for other factors (aOR: 0.45; 95% CI [0.25 – 0.81]).

**Conclusion:** An early short course of methylprednisolone in patients with moderate to severe COVID-19 reduced escalation of care and improved clinical outcomes.

**Summary:** In this multi-center quasi-experimental study of 213 patients, we demonstrate early short course of methylprednisolone in moderate to severe COVID-19 patients reduced the composite endpoint of escalation of care from ward to ICU, new requirement for mechanical ventilation, and mortality.

## Background

As of April 9th, 2020, the United States has over 400,000 cases of confirmed coronavirus disease 2019 (COVID-19) caused by the severe acute respiratory syndrome coronavirus 2 (SARS-CoV-2) [1]. Most patients will have mild illness, but older persons and those with comorbidities may develop severe disease necessitating hospitalization and intensive unit (ICU) care [2, 3]. The disease pathophysiology presents in two distinct overlapping phases, the initial pathogenic viral response followed by host inflammatory response with grades of severity associated with distinct clinical findings [4,5]. The pathological progression in severe COVID-19 includes an excessive and unregulated pro-inflammatory cytokine storm resulting in immunopathological lung injury, diffuse alveolar damage with the development of acute respiratory distress syndrome (ARDS), and death [6–9].

In the absence of any proven anti-viral therapy the current clinical management is primarily supportive care, supplemental oxygen, and mechanical ventilatory support [1,10]. Adjunctive therapy with immunomodulatory agents targeting the inflammatory cytokine storm are being evaluated [5,10]. Studies of corticosteroid therapy for phylogenetically similar coronavirus infections showed no benefit and potential harm [11]. Despite the frequent use in treating patients with COVID-19 in China, the role of corticosteroids is undefined [3,5,10–13]. A more recent observational study reported improved outcomes in patients with COVID-associated ARDS that received corticosteroids [14].

We postulated early treatment with a short course of corticosteroids in patients with COVID-19 may attenuate the excessive host respiratory and systemic inflammatory responses. We report the clinical characteristics and early outcomes of patients with COVID-19 receiving short courses of methylprednisolone.

## Methods

### Study Population

Consecutive patients hospitalized between March 12, 2020 through March 27, 2020 were eligible for inclusion if they were 18 years of age or older, had confirmed COVID-19 infection, with radiographic evidence of bilateral pulmonary infiltrates, and required oxygen by nasal cannula, high-flow nasal cannula (HFNC), or mechanical ventilation. Patients were excluded if they were transferred from an out-of-system hospital, died within 24 hours of presentation to the ED, or were admitted for less than 24 hours. A confirmed case of COVID-19 was defined as a patient that had a positive reverse-transcriptase–polymerase- chain-reaction (RT-PCR) assay for SARS-CoV-2 in a nasopharyngeal sample tested by the Michigan Department of Health and Human Services (MDHHS) or the Henry Ford Health System (HFHS) centralized clinical microbiology laboratory. Beginning March 16, 2020, testing for hospitalized patients was performed by the centralized clinical microbiology laboratory.

Patients were risk stratified by severity of symptoms on presentation to the hospital as mild, moderate, or severe COVID-19. Patients without hypoxia or exertional dyspnea were considered to have mild COVID-19. Patients with mild COVID-19 were treated with symptom relief only and not admitted to the hospital. Patients who presented with infiltrates on chest radiography and required supplemental oxygen by nasal cannula or HFNC were classified as having moderate COVID-19. Patients who had respiratory failure requiring mechanical ventilation were classified as having severe COVID-19.

### Study Design

This was a multi-center quasi-experimental study at HFHS, comprised of five hospitals in southeast and south-central Michigan. The study was approved by the institution’s Investigational Review Board (#13739) with waiver of consent. Patients in the pre-corticosteroid protocol group from March 12, 2020 through March 19, 2020 were compared to a corticosteroid protocol group that included patients from March 20, 2020 through March 27, 2020.

Patients in both study groups received standard care, comprised of supplemental oxygen, HFNC, invasive ventilation, antibiotic agents, antiviral agents, vasopressor support, and renal-replacement therapy, as determined by the primary team. Patients who progressed to ARDS were managed with standard of care [15].

## Intervention

### Pre-Corticosteroid Protocol

Patients with moderate or severe disease who presented to HFHS within the first week of the COVID epidemic in Detroit were initially treated with supportive care with or without a combination of lopinavir-ritonavir and ribavirin or hydroxychloroquine according an institutional guideline developed by Infectious Diseases Physicians and Pharmacists. The institutional guidelines were developed by consensus, and based on the available literature, experience from Wuhan, China and other centers around the world affected by COVID-19 before Michigan. Intravenous (IV) remdesivir compassionate use was requested for eligible mechanically ventilated patients. On March 17, 2020 lopinavir-ritonavir with ribavirin was removed from the COVID-19 institutional protocol.[16]

### Corticosteroid Protocol

As a result of observed poor outcomes, clinical rationale based upon immunology, clinical course of COVID-19, and more recently best available evidence, the HFHS corticosteroid protocol was developed (Supplementary Materials) [14,15,17]. We hypothesized that early corticosteroids would combat the inflammatory cascade leading to respiratory failure, ICU escalation of care, and mechanical ventilation. The corticosteroid protocol became the institutional standard on March 20, 2020. Patients with confirmed influenza infection were not recommended to receive corticosteroids.

Moderate COVID-19 was treated with hydroxychloroquine 400 mg twice daily for 2 doses on day 1, followed by 200 mg twice daily on days 2-5. Patients with moderate COVID-19 who required 4 liters or more of oxygen per minute on admission, or who had escalating oxygen requirements from baseline, were recommended to receive IV methylprednisolone 0.5 to 1 mg/kg/day in 2 divided doses for 3 days. Patients who required ICU admission were recommended to receive the above regimen of hydroxychloroquine and IV methylprednisolone 0.5 to 1 mg/kg/day in 2 divided doses for 3 to 7 days. ICU patients were also evaluated for tocilizumab on a case-by-case basis. Oral switch was performed to prednisone at a ratio of 1 to 1 when determined clinically appropriate by the primary medical team.

## Data Collection

Data was ascertained from each institution’s electronic medical record and recorded in a standardized electronic case report form. Demographic data, information on clinical symptoms or signs at presentation, and laboratory and radiologic results during admission. All laboratory tests and radiologic assessments, including plain chest radiography and computed tomography of the chest, were performed at the discretion of the treating physician.

## Study Definitions

We ascertained coexisting conditions from electronic medical record and physician documentation. The National Early Warning Score (NEWS) was collected to evaluate baseline illness severity based on vital signs obtained in the Emergency Department [18]. Additionally, the quick Sequential Organ Failure Assessment (qSOFA) was used to evaluate severity of illness of included patients based on ED vitals and examination [19]. All patients were followed for at least 14 days after initial presentation. Patient data was censored on April 9, 2020.

### Outcome Measures

#### Primary Endpoint

The primary composite endpoint was escalation to intensive care unit (ICU) from a general medical unit (GMU), progression to respiratory failure requiring mechanical ventilation after hospital admission, or in-hospital all-cause mortality. Patients directly admitted to the ICU from the emergency room were evaluated for the latter two outcomes and those requiring mechanical ventilation in the emergency room were evaluated for mortality.

#### Secondary Endpoints

Secondary endpoints included development and severity of ARDS, days to ventilator liberation, shock, acute kidney injury (AKI), and length of hospital stay (LOS). ARDS was diagnosed and classified according to the Berlin Definition [20]. AKI was diagnosed according to the Kidney Disease: Improving Global Outcomes definition [21].

### Statistical Analysis

Continuous variables were reported as median and interquartile range (IQR) and compared using the Mann-Whitney test or t-test, as appropriate. Categorical data was reported as number and percentage (no., %) and compared using the chi-squared test or Fisher’s exact test, as appropriate. No imputation was made for missing data points. The sample size was derived from all eligible consecutive hospitalized patients during the study period. A two-sided α < 0.05 was considered statistically significant. Bivariate and multivariable logistic regression analysis was planned a-priori to test the association between the composite endpoint and exposure to the corticosteroid protocol. Covariates in the bivariate analysis with a p-value <0.2 and clinical rationale were included in a multivariable regression model that was restricted to a subject-to-variable ratio of 10:1. To evaluate the implementation timing of the in-house RT-PCR SARSCoV-2 testing a post-hoc sensitivity analysis were conducted on the composite outcome. A non-equivalent dependent variable, receipt of, and time to empiric antibiotic therapy for pneumonia, was utilized to account for potential maturation in the management of COVID-19. Statistical analysis was performed using IBM SPSS version 25 (Chicago, IL) and SAS 9.4 (Cary, NC).

## Results

Two-hundred and fifty consecutive patients were evaluated for inclusion. Ten were hospitalized for 24 hours or less, 23 did not require oxygen by nasal cannula, HFNC or mechanical ventilation, and four expired within 24 hours of admission. Two-hundred and thirteen patients were included, 81 (38%) in the pre-corticosteroid protocol group and 132 (62%) in the corticosteroid protocol group. The median age of the pre-corticosteroid protocol group and corticosteroid protocol group was 64 (IQR: 51.5, 73.5) and 61 (IQR: 51, 72) years, respectively. Black patients comprised 61.7% of the pre-corticosteroid protocol group and 79.5% of the corticosteroid protocol group (p=0.005). Of the comorbid conditions evaluated, chronic obstructive pulmonary disease was more frequent in the pre-corticosteroid protocol group compared to the corticosteroid protocol group (18.5% vs 9.1%; p=0.045). The presenting COVID-19 symptoms, baseline severity of illness, and other demographics were similar between groups (Table I).

**Table I.**
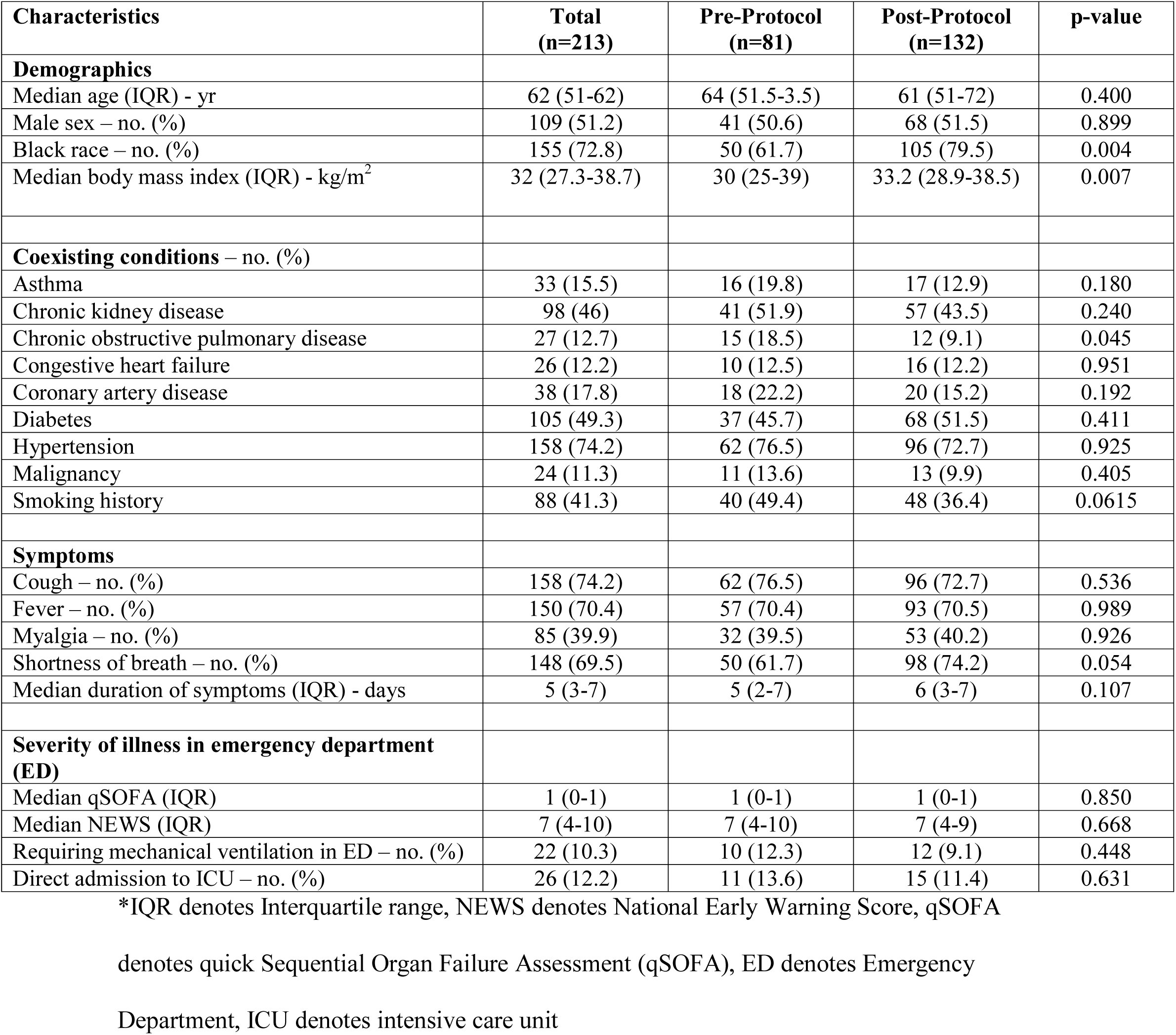
Baseline Demographics and Clinical Characteristics of Study Patients.

| Characteristics | Total<br>(n=213) | Pre-Protocol<br>(n=81) | Post-Protocol<br>(n=132) | p-value |
| --- | --- | --- | --- | --- |
| **Demographics** |  |  |  |  |
| Median age (IQR) - yr | 62 (51-62) | 64 (51.5-3.5) | 61 (51-72) | 0.400 |
| Male sex – no. (%) | 109 (51.2) | 41 (50.6) | 68 (51.5) | 0.899 |
| Black race – no. (%) | 155 (72.8) | 50 (61.7) | 105 (79.5) | 0.004 |
| Median body mass index (IQR) - kg/m<sup>2</sup> | 32 (27.3-38.7) | 30 (25-39) | 33.2 (28.9-38.5) | 0.007 |
| **Coexisting conditions – no. (%)** |  |  |  |  |
| Asthma | 33 (15.5) | 16 (19.8) | 17 (12.9) | 0.180 |
| Chronic kidney disease | 98 (46) | 41 (51.9) | 57 (43.5) | 0.240 |
| Chronic obstructive pulmonary disease | 27 (12.7) | 15 (18.5) | 12 (9.1) | 0.045 |
| Congestive heart failure | 26 (12.2) | 10 (12.5) | 16 (12.2) | 0.951 |
| Coronary artery disease | 38 (17.8) | 18 (22.2) | 20 (15.2) | 0.192 |
| Diabetes | 105 (49.3) | 37 (45.7) | 68 (51.5) | 0.411 |
| Hypertension | 158 (74.2) | 62 (76.5) | 96 (72.7) | 0.925 |
| Malignancy | 24 (11.3) | 11 (13.6) | 13 (9.9) | 0.405 |
| Smoking history | 88 (41.3) | 40 (49.4) | 48 (36.4) | 0.0615 |
| **Symptoms** |  |  |  |  |
| Cough – no. (%) | 158 (74.2) | 62 (76.5) | 96 (72.7) | 0.536 |
| Fever – no. (%) | 150 (70.4) | 57 (70.4) | 93 (70.5) | 0.989 |
| Myalgia – no. (%) | 85 (39.9) | 32 (39.5) | 53 (40.2) | 0.926 |
| Shortness of breath – no. (%) | 148 (69.5) | 50 (61.7) | 98 (74.2) | 0.054 |
| Median duration of symptoms (IQR) - days | 5 (3-7) | 5 (2-7) | 6 (3-7) | 0.107 |
| **Severity of illness in emergency department (ED)** |  |  |  |  |
| Median qSOFA (IQR) | 1 (0-1) | 1 (0-1) | 1 (0-1) | 0.850 |
| Median NEWS (IQR) | 7 (4-10) | 7 (4-10) | 7 (4-9) | 0.668 |
| Requiring mechanical ventilation in ED – no. (%) | 22 (10.3) | 10 (12.3) | 12 (9.1) | 0.448 |
| Direct admission to ICU – no. (%) | 26 (12.2) | 11 (13.6) | 15 (11.4) | 0.631 |

\*IQR denotes Interquartile range, NEWS denotes National Early Warning Score, qSOFA
denotes quick Sequential Organ Failure Assessment (qSOFA), ED denotes Emergency
Department, ICU denotes intensive care unit

Overall, corticosteroids use was 56.8% and 68.2% in the pre-corticosteroid protocol group and corticosteroid protocol group, respectively (p=0.094). The corticosteroid protocol group had a greater proportion of corticosteroids initiated within 48 hours of presentation (12.4% vs. 41.7%, p < 0.001). The median time to corticosteroid initiation was 5 days (IQR 3-7, range 1-9) in the pre-corticosteroid protocol group and 2 days (IQR 1-3, range 0-8) in the corticosteroid protocol group. The median time to hydroxychloroquine initiation was earlier in the precorticosteroid protocol group compared to the corticosteroid protocol group (3 [IQR: 1, 4] vs. 1 [IQR: 0, 2] days, p < 0.001). Additional treatment characteristics are described in Table II.

**Table II.**
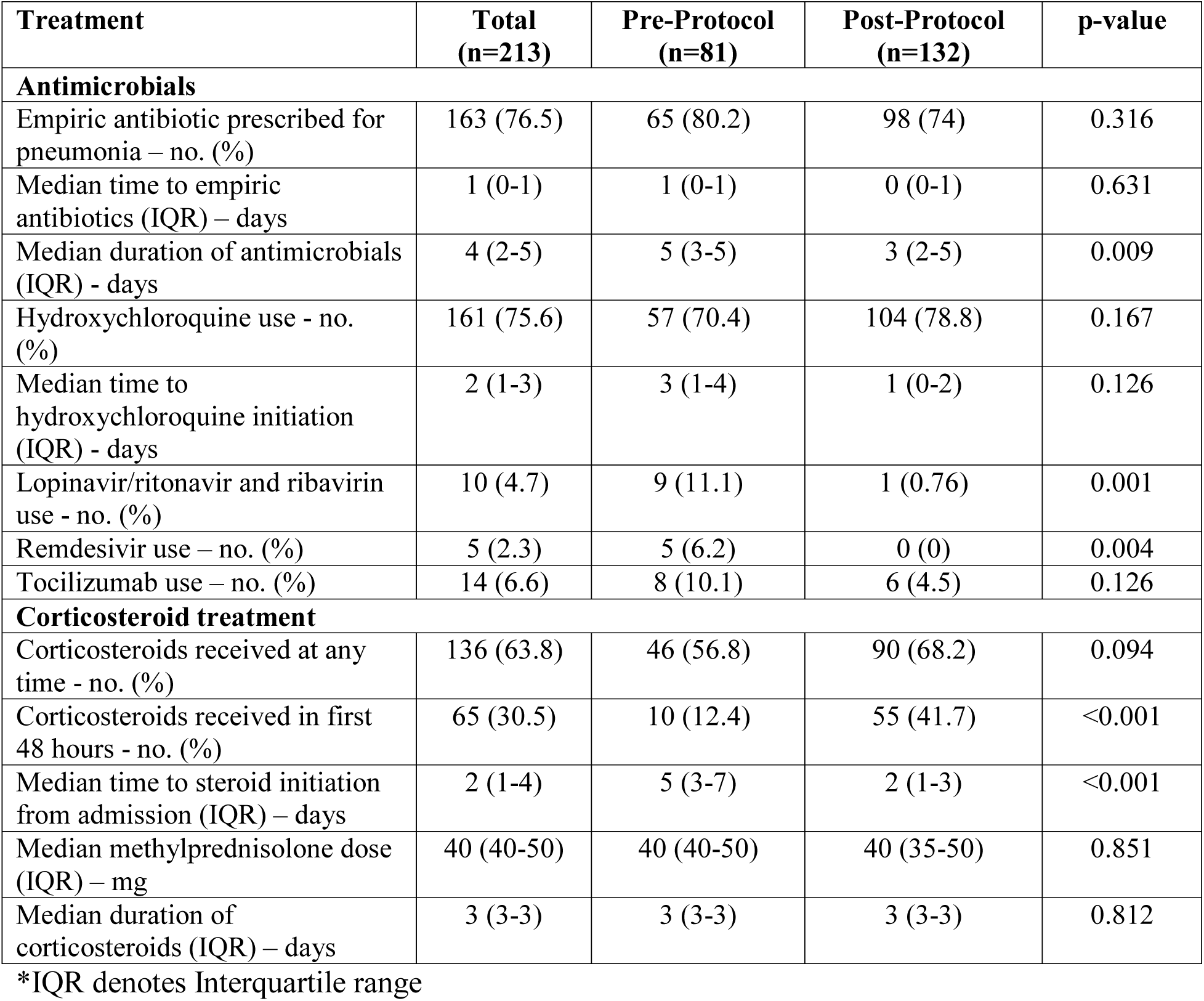
Treatments Received by Patients Pre-Corticosteroid and Corticosteroid Protocol Groups.

| Treatment | Total<br>(n=213) | Pre-Protocol<br>(n=81) | Post-Protocol<br>(n=132) | p-value |
| --- | --- | --- | --- | --- |
| **Antimicrobials** |  |  |  |  |
| Empiric antibiotic prescribed for pneumonia – no. (%) | 163 (76.5) | 65 (80.2) | 98 (74) | 0.316 |
| Median time to empiric antibiotics (IQR) – days | 1 (0-1) | 1 (0-1) | 0 (0-1) | 0.631 |
| Median duration of antimicrobials (IQR) - days | 4 (2-5) | 5 (3-5) | 3 (2-5) | 0.009 |
| Hydroxychloroquine use - no. (%) | 161 (75.6) | 57 (70.4) | 104 (78.8) | 0.167 |
| Median time to hydroxychloroquine initiation (IQR) - days | 2 (1-3) | 3 (1-4) | 1 (0-2) | 0.126 |
| Lopinavir/ritonavir and ribavirin use - no. (%) | 10 (4.7) | 9 (11.1) | 1 (0.76) | 0.001 |
| Remdesivir use – no. (%) | 5 (2.3) | 5 (6.2) | 0 (0) | 0.004 |
| Tocilizumab use – no. (%) | 14 (6.6) | 8 (10.1) | 6 (4.5) | 0.126 |
| **Corticosteroid treatment** |  |  |  |  |
| Corticosteroids received at any time - no. (%) | 136 (63.8) | 46 (56.8) | 90 (68.2) | 0.094 |
| Corticosteroids received in first 48 hours - no. (%) | 65 (30.5) | 10 (12.4) | 55 (41.7) | <0.001 |
| Median time to steroid initiation from admission (IQR) – days | 2 (1-4) | 5 (3-7) | 2 (1-3) | <0.001 |
| Median methylprednisolone dose (IQR) – mg | 40 (40-50) | 40 (40-50) | 40 (35-50) | 0.851 |
| Median duration of corticosteroids (IQR) – days | 3 (3-3) | 3 (3-3) | 3 (3-3) | 0.812 |

\*IQR denotes Interquartile range

The primary composite endpoint occurred at a significantly lower rate in corticosteroid protocol group compared to the pre-corticosteroid protocol group (34.9% vs. 54.3%, p=0.005). A significant reduction in each of primary composite endpoints was also noted. In the sensitivity analysis sub-group, after the implementation of the in-house RT-PCR SARS-CoV-2 testing, 34.9% (46 of 132 patients) and 55% (33 of 60 patients) experienced the primary composite endpoint in the corticosteroid protocol group and pre-corticosteroid protocol group, respectively (p=0.009).

The median LOS was significantly reduced from 8 to 5 days in pre-corticosteroid protocol group compared to the corticosteroid protocol group (p < 0.001). ARDS occurred in 38.3% and 26.6% in the pre-corticosteroid protocol group and corticosteroid protocol group, respectively (P=0.04). After adjustment for male sex and age over 60, the corticosteroid protocol was independently associated with a reduction in the composite endpoint at 14 days (aOR: 0.45; 95% CI [0.25 – 0.81]) (Table 2, Supplemental Materials). Outcomes at 14 days also included 9 (11.1%) of pre-steroid patients remaining admitted as compared to 26 (19.7%) of post-steroid patients. Table III describes additional outcomes before and after implementation of the corticosteroid protocol.

**Table III.** Outcomes in the Pre-Corticosteroid and Corticosteroid Protocol Groups.

|  | Pre-Protocol<br>(n=81) | Post-Protocol<br>(n=132) | Odds Ratio (CI) | p-value |
| --- | --- | --- | --- | --- |
| **Primary Outcome** |  |  |  |  |
| Primary composite outcome – no. (%) | 44 (54.3) | 46 (34.9) | 0.45 (0.26 – 0.79) | 0.005 |
| Death – no. (%) | 21 (26.3) | 18 (13.6) | 0.45 (0.22 – 0.91) | 0.024 |
| Respiratory failure requiring mechanical ventilation – no. (%)* | 26 (36.6) | 26 (21.7) | 0.47 (0.25-0.92) | 0.025 |
| Escalation from GMU to ICU – no. (%)<sup>+</sup> | 31 (44.3) | 32 (27.3) | 0.47 (0.25 – 0.88) | 0.017 |
| **Secondary Outcomes** |  |  |  |  |
| Overall mechanical ventilation – no. (%) | 36 (44.4) | 38 (28.8) | 0.51 (0.28 – 0.90) | 0.020 |
| ARDS – no. (%) | 31 (38.3) | 33 (26.6) |  | 0.040 |
| Mild | 3 (3.7) | 1 (0.76) |  | 0.125 |
| Moderate | 8 (9.9) | 9 (6.8) |  | 0.307 |
| Severe | 20 (24.7) | 23 (17.4) |  | 0.201 |
| Median duration of mechanical ventilation (IQR) - days | 8 (4-13) | 7 (4-9) |  | 0.558 |
| Median time to extubation (IQR) – days | 8 (4-13) | 7 (4-9) |  | 0.558 |
| Shock – no. (%) | 19 (23.5) | 17 (12.6) |  | 0.069 |
| Acute kidney injury – no. (%) | 42 (51.9) | 59 (44.7) |  | 0.310 |
| Median hospital length of stay (IQR) - days | 8 (5-14) | 5 (3-7) |  | <0.001 |
| Discharged from hospital – no. (%) | 51 (62.2) | 88 (66.7) |  | 0.584 |
| Remain hospitalized – no. (%) | 9 (11.1) | 26 (19.7) |  | 0.102 |
| Remain intubated – no. (%) | 7 (8.6) | 13 (9.8) |  | 0.771 |

CI denotes confidence interval, ICU denotes intensive care unit, GMU denotes general medical unit, ARDS denotes acute respiratory distress syndrome, IQR denotes Interquartile range *A total of 10 and 12 patients were not included in this analysis because they required mechanical ventilation in the emergency department in the pre-corticosteroid and corticosteroid group, respectively.

+A total of 11 and 15 patients were not included in this analysis because they were directly admitted to the intensive care unit in the pre-corticosteroid and corticosteroid group, respectively.

## Discussion

In this quasi-experimental study, hospitalized patients with moderate to severe COVID-19 that received an early short course of methylprednisolone had a reduced rate of the primary composite endpoint of death, ICU transfer, and mechanical ventilation, with a number needed to treat of 8 to prevent one patient transfer for mechanical ventilation. The reduction in ICU transfer and requirement for mechanical ventilation represents a potential intervention to reduce critical care utilization during the COVID-19 pandemic [22,23]. The median reduction of hospital LOS by three days observed with the use of corticosteroids, could positively impact hospital capacity during the COVID-19 surge.

Corticosteroids are not routinely recommended in patients with COVID-19 without an alternate indication or presence of ARDS [11,15,24]. Data are conflicting; corticosteroid use in previous viral respiratory illnesses have demonstrated delayed viral clearance and increased mortality [11,25]. On the contrary, short course corticosteroids in some reports are beneficial and safe in critically ill patients with SARS-CoV-2 and were not found to be an independent risk factor of prolonged viral RNA shedding [17,26,27]. These discordant findings may be explained by the observational nature of the studies, heterogeneity in patient acuity, inconsistent dosing regimens and duration, and timing of initiation of therapy [10,17]. Corticosteroids were used in 11-35% of non-severe and 45–72% of severe COVID-19 cases in China, however the benefits and risks remain undefined [2,6,12–14]. A mortality benefit (HR, 0.38: 95% CI, 0.20–0.72) with the use methylprednisolone was reported in one retrospective cohort study of COVID-19 patients with ARDS [14].

COVID-19 can progress from mild to severe illness characterized by an initial viral infection phase followed by pulmonary inflammation, and then a hyper-inflammation phase [4,6,9]. The pulmonary phase is associated with progressive dyspnea and radiographic findings of pneumonia.[4] Symptom onset to dyspnea and ARDS development occurs between a median of 5 to 7 days and 8 to 12 days, respectively [6,12,28]. The present study findings support that a short course of methylprednisolone may attenuate progression to the hyper-inflammation phase that requires escalation of care in patients with COVID-19. Hydroxychloroquine with or without azithromycin, remdesivir, and lopinavir/ritonavir with ribavirin were prescribed at similar frequency between groups. These agents have demonstrated mixed efficacy results for COVID-19 in placebo controlled-trials, with hydroxychloroquine being the most promising at this time [10]. Immunomodulatory agents, such as tocilizumab, were infrequently used in this study.

This study has several limitations. Given the pandemic nature of the disease a pragmatic quasi-experimental design was used. The potential for regression to the mean and maturation is an inherent limitation to all quasi-experiments. On March 16, 2020 rapid on-site RT-PCR testing for SARS-CoV-2 testing was implemented, and some of the pre-steroid group experienced delayed diagnosis and treatment. However, the observed association was unchanged in the sensitivity analysis. A non-equivalent dependent variable, empiric antibiotic therapy for pneumonia, suggested no difference in management of COVID-19. Some of the pre-corticosteroid protocol group received corticosteroids after initiation of the updated COVID-19 institutional treatment protocol. Steroids initiated in this group were started significantly later. Additionally, guideline adherence in the corticosteroid protocol group was not universal. Finally, the study has a limited follow up period of 14 days, similar to other recent reports. As of April 9, 2020, 51 (62.9%) of patients in the pre-steroid cohort and 88 (66.7%) of patients in the post-steroid cohort were discharged from the hospital. As a result, outcomes for those patients are not known. Anecdotally, we observed hyperglycemia, but no severe corticosteroid related adverse effects (i.e. gastrointestinal hemorrhage), and data collection is ongoing.

In conclusion, early use of a short course of methylprednisolone, an inexpensive and readily available agent, in patients with moderate to severe COVID-19 may prevent progression of disease and improve outcomes. These findings are crucial given the ongoing COVID-19 pandemic and ICU bed and mechanical ventilator shortages. Research is urgently needed to further define the role of corticosteroids in patients with COVID-19 at a high-risk of clinical deterioration, identified early in the disease course using prognostic markers or clinical prediction tools.

## Data Availability

Data will be available after one year from publication.

## Acknowledgements

^#^Henry Ford COVID-19 Task Force

Varidhi Nauriyal, M.D.^1,2^, Jayanth Lakshmikanth, M.D.^2^, Asif Abdul Hamed, M.D.^2^, Owais Nadeem, M.D.^2^, Kristin Griebe, Pharm.D. ^3^, Joseph M. Johnson, Pharm.D. ^3^, Patrick Bradley, M.D.^2^, Junior Uduman, M.D.^2^, Sara Hegab, M.D.^2^, Jennifer Swiderek, M.D.^2^, Amanda Godfrey, M.D.^2^, Jeffrey Jennings, M.D.^2^, Jayna Gardner-Gray, M.D.^5^, Adam Ackerman, M.D.^6^, Jonathan Lezotte, M.D.^6^, Joseph Ruhala, M.D.^6^, Linoj Samuel, PhD, D(ABMM).^7^, Robert J. Tibbetts, Ph.D. D(ABMM) F(CCM).^7^, Indira Brar, M.D.^1^, John McKinnon, M.D.^1^, Geehan Suleyman, M.D.^1^, Nicholas Yared, M.D.^1^, Erica Herc, M.D.^1^, Jonathan Williams, M.D.^1^, Odaliz Abreu Lanfranco, M.D.^1^, Anne Chen, M.D.^1^, Marcus Zervos, M.D.^1^

Eric Scher, M.D.^4^, for his courageous leadership and commitment to education in the field of Academic Internal Medicine. Krishna Modi, M.D.^4^, Rebecca Bussa, Kelly Curran, M.D.^4^, Abigail Entz, M.D.^4^, Hafsa Abdulla, M.D.^4^, and Charles Hammond, M.D.^4^, for data collection and review. We thank the entire front-line staff members of the Henry Ford Health System for dedicated and compassionate patient care during the COVID-19 outbreak.

^1^Infectious Diseases, Henry Ford Hospital, 2799 West Grand Blvd, Detroit, MI, 48202;

^2^Pulmonary Medicine, Henry Ford Hospital, 2799 West Grand Blvd, Detroit, MI, 48202

^3^Pharmacy, Henry Ford Hospital, 2799 West Grand Blvd, Detroit, MI, 48202;

^4^Internal Medicine, Henry Ford Hospital, 2799 West Grand Blvd, Detroit, MI, 48202

^5^Emergency Medicine, Henry Ford Hospital, 2799 West Grand Blvd, Detroit, MI, 48202

^6^Surgical Critical Care, Henry Ford Hospital, 2799 West Grand Blvd, Detroit, MI, 48202

^7^Pathology and Microbiology, Henry Ford Hospital, 2799 West Grand Blvd, Detroit, MI, 48202

---

**Figure 1.**
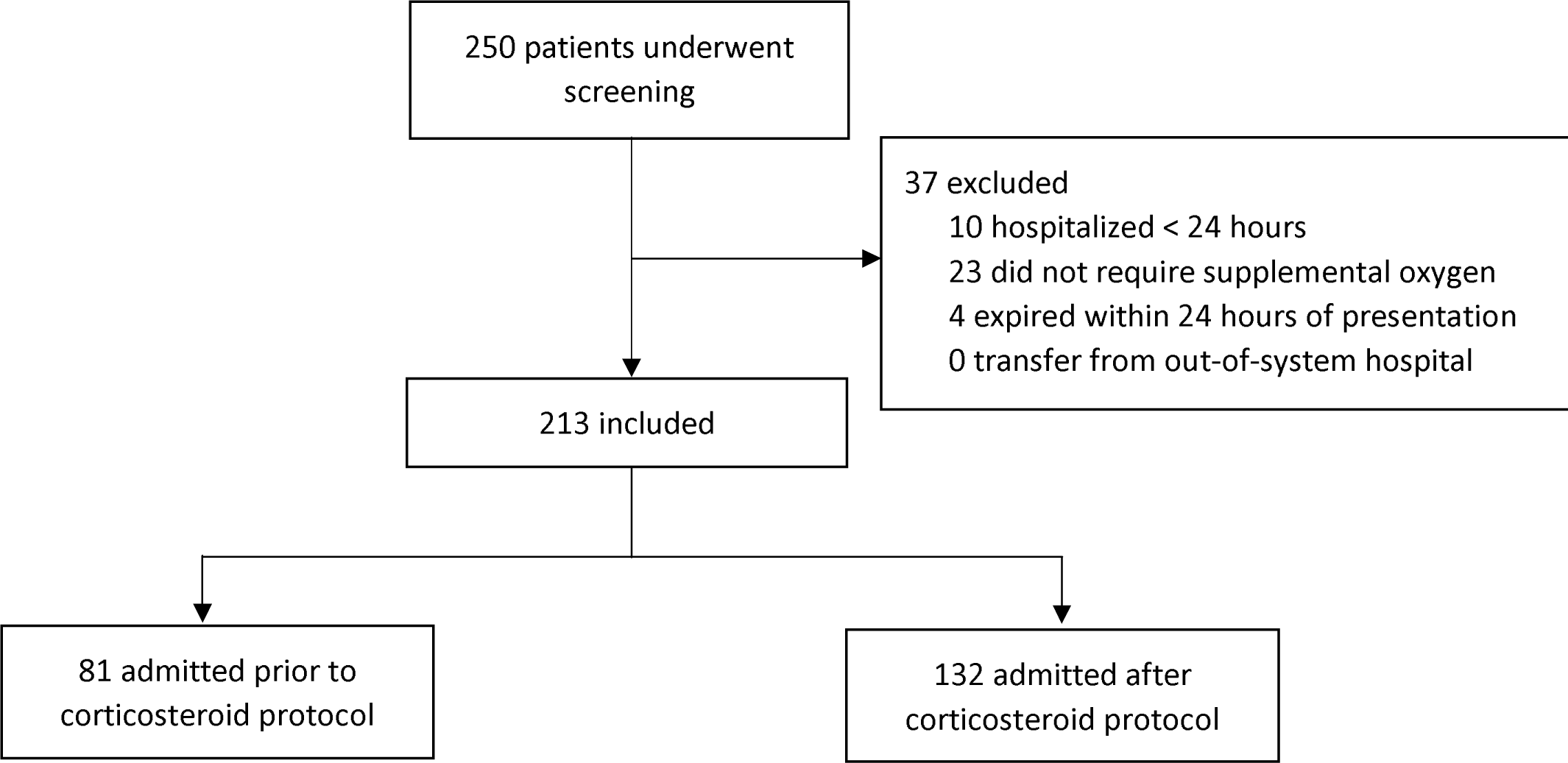
Number of Patients Screened and Included in the Trial.

**Table 1.**
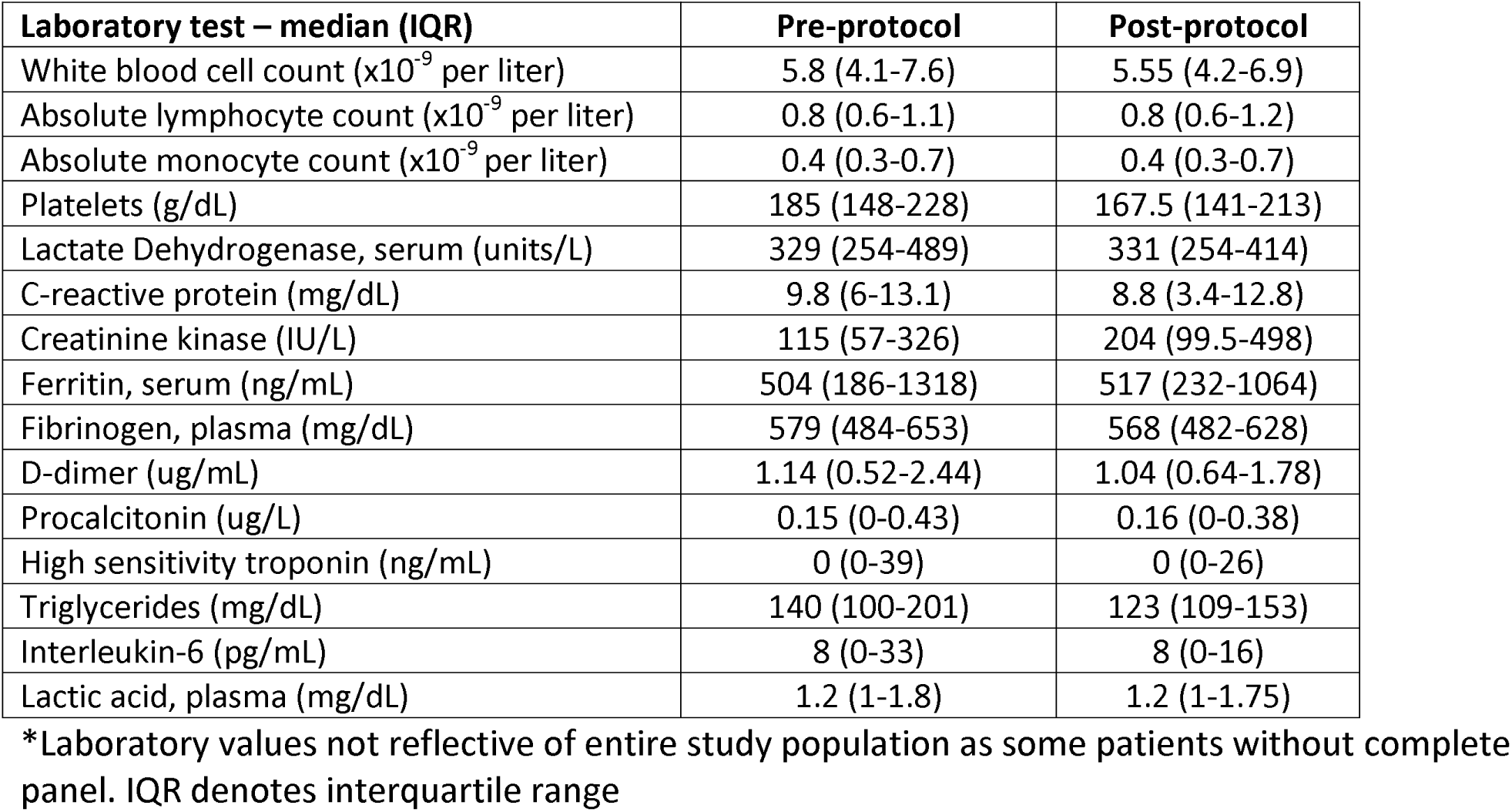
Baseline laboratory tests for the pre- and post-corticosteroid protocol populations^*^.

**Table 2.** Multivariate Logistic Regression for Effect on Composite Endpoint at 14 Days.

| Covariate | Adjusted odds ratio | p-value |
| --- | --- | --- |
| Corticosteroid protocol | 0.45 (0.25 – 0.81) | 0.007 |
| Male gender | 2.20 (1.23 – 3.91) | 0.008 |
| Age greater than 60 (years) | 1.97 (1.1 – 3.54) | 0.022 |

**Figure 1.**
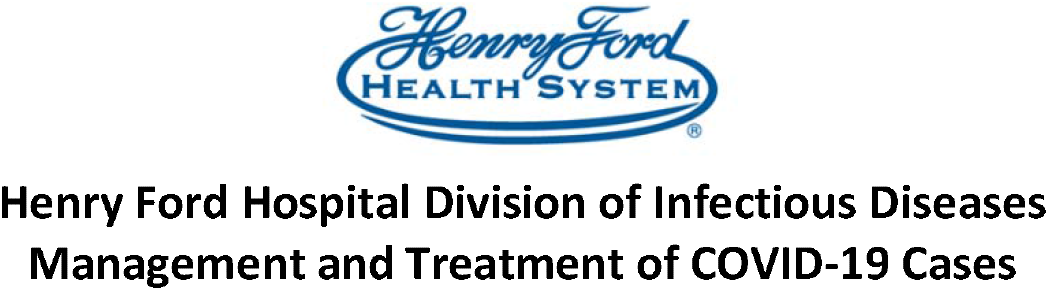
Management and Treatment of COVID-19 Cases Guideline.

All confirmed COVID-19 inpatients require Infectious Diseases consultation for management.

For testing recommendations, please refer to SARS-CoV-2 (COVID-19) Testing Criteria from HFHS infection prevention and control.

Currently, there are <u>no FDA approved therapies to treat COVID-19</u>. Some medications have demonstrated in vitro, animal, or very limited clinical safety and efficacy data in other coronaviruses (e.g. SARS and MERS). The Division of Infectious Diseases is coordinating research/compassionate use remdesivir. The below treatments should be started immediately once COVID test is positive (when indicated), pending remdesivir availability. **Preemptively starting COVID medications prior to return of test results is not recommended due to limited medication supply unless there is high suspicion based upon clinical judgement and patient characteristics**. These guidelines are interim recommendations and may change according to drug availability and new data published. Supportive care and infection control measures are indicated for all hospitalized patients.

**Initial labs for all inpatients** (see below for ongoing treatment monitoring):

- <u>Suspected or confirmed</u> patients: CBC with differential, BMP, magnesium, ferritin, liver profile, bilirubin, total, procalcitonin, CPK, D-dimer, CRP, LDH, high sensitivity troponin
- Draw upon admission to **ICU:** Triglyceride, IL-6, DIC panel (in addition to labs listed above if not previously ordered)
- Baseline EKG

| Treatment for COVID Patients |  |  |  |
| --- | --- | --- | --- |
|  | Mild symptoms, outpatient | Moderate symptoms, hospitalized | Severe, hospitalized |
| Characteristics | Cough, fever, myalgias<br>On Room air / baseline SAO<sub>2</sub> 94% or above<br>No radiographic evidence of pulmonary infiltrates | **Supplemental oxygen use or Sao<sub>2</sub> less than 94%**, fever of ≥ 36.6°C armpit, ≥ 37.2 °C oral, or ≥ 37.8°C rectal, radiographic evidence of pulmonary infiltrates | Respiratory failure requiring **mechanical ventilation**<br>Cough, fever, myalgias |
| Recommended treatment | Social distancing recommended<br><u>Consider OTC Symptom relief</u><br>Cough:<br>-vaporizer or humidifier<br>-dextromethorphan (dry | **\*Hydroxychloroquine** 400 mg PO BID x 2 doses, then 200 mg PO BID x 4 days<br>OR<br>**Remdesivir**: Requires patient | **\*Hydroxychloroquine** 400mg PO BID x 2 doses, then 200 mg PO BID x 4 days<br>OR<br>**Remdesivir**: Expanded access |
|  | cough)<br>-guaifenesin (productive cough)<br>Fever:<br>Acetaminophen | informed consent for clinical trial. Not an FDA approved therapy | Requires patient informed consent. Not an FDA approved therapy |

| Treatment for COVID Patients (continued) |  |  |  |
| --- | --- | --- | --- |
|  | Mild symptoms, outpatient | Moderate symptoms, hospitalized | Severe, hospitalized |
| Adjunctive Therapy |  | - Steroids routinely **not** recommended unless indicated for another comorbidity (e.g. COPD)<br>**New supplemental oxygen $\geq 4$ L or escalating oxygen requirements from baseline**<br>- Recommend **low dose methylprednisolone** with a short course (e.g. 0.5-1 mg/kg/day based on actual body weight divided in 2 doses for 3 days)<br>- If severe hypoxia with continued supplemental oxygen requirement on day 3, consider extending methylprednisolone to a total duration of 5 to 7 days | - Steroids routinely **not** recommended unless indicated for another comorbidity (e.g. COPD)<br>**Mechanical ventilation**<br>- Recommend **low dose methylprednisolone** with a short course (e.g. 0.5-1 mg/kg/day based on actual body weight divided in 2 doses for 3 days)<br>- If severe hypoxia with continued supplemental oxygen requirement on day 3, consider extending methylprednisolone to a total duration of 5 to 7 days<br>Additional immunomodulator usage will be determined on a case-by-case basis with the ICU and ID COVID team |

**NOTES**: If **influenza** testing was done and is **positive, steroids are not recommended**.

#Consider a dose maximum of methylprednisolone 80 mg IV every 12 hours for obese patients. When clinically appropriate for oral switch, convert methylprednisolone 1:1 to oral prednisone.

Monitoring Parameters
- Hydroxychloroquine: Cardiotoxicity, Torsade de Pointes, depression, psychosis. Maintain potassium at least 4 mEq/L, and magnesium at least 2 mEq/L. **Refer to QT monitoring appendix**.
- Remdesivir: Phlebitis, Constipation, Nausea, Headache, Bruising, Liver Function test abnormalities. Obtain daily BMP and LFTs.

